# LLM-based data extraction for a large cancer registry, the Ontario Hereditary Cancer Research Network

**DOI:** 10.1101/2025.08.20.25334127

**Authors:** Andres Felipe Melani De La Hoz, Jochen Weile, Pratham Hemlani, Elif Tuzlali, Sarah Ridd, Brandon Chan, Lauren K. Hughes, Kathy Chun, Harriet Feilotter, Daria Grafodatskaya, Jordan Lerner-Ellis, Laila Schenkel, Amanda Smith, Andrea Vaags, Hong Wang, Raymond H. Kim, Lincoln Stein, Benjamin Haibe-Kains, Melanie Courtot

**Author notes:** These authors contributed equally to this work (Andres Felipe Melani De La Hoz, Jochen Weile, Pratham Hemlani).

## Abstract

**Importance:** Manual data extraction from genomic lab reports for on-line registries and databases is time-consuming for human resources such as clinical research coordinators. Automated tools, especially LLMs, can address these issues. Efficient and accurate data processing is crucial for building a reliable database.

**Objective:** To streamline the data extraction and curation process for genetic testing lab reports using an LLM-based approach.

**Design:** Nine sample molecular lab reports were selected for manual data extraction by two expert curators. The process was timed, and the results served as gold-standard for validating automated extraction. Eighteen fields from the OHRCN’s data model were selected as extraction targets.

**Setting:** The study was conducted within OHCRN, which unifies research, genomic, and clinical patient data from clinics and laboratories across Ontario, Canada.

**Participants:** Nine laboratories agreed to share sample molecular lab reports and two clinical research coordinators affiliated with OHCRN participated as data curators.

**Exposure:** LLM-based Extraction of Information (LEI), an automated data extraction pipeline, was developed using regular expressions, Trie search, and LLMs to extract data from molecular lab reports and structure it for inclusion into OHCRN’s database.

**Main Outcomes and Measures:** LEI was evaluated by measuring the F1-score on the extraction task of 18 entity types. These measures were compared against 15 extraction tools in the biomedical domain. Extraction time was also measured and compared against manual extraction times.

**Results:** LEI demonstrated quality on par with and surpassing other existing LLM-based extraction methods. Reference tools showed F1-scores around 70%, while LEI achieved an average score of 87.4%. LEI reduced extraction time by approximately 2-fold, with an average time of 7.59 minutes per report including results review by curators, compared to 14.88 minutes per report for manual extraction.

**Conclusions and Relevance:** LEI facilitates standardized, accurate, and efficient healthcare data extraction from unstructured texts, significantly improving the current OHCRN workflow. By automating the extraction process, LEI allows expert curators to focus on validating results rather than performing manual data entry. LEI’s simple interface enables researchers to easily guide extraction tasks and supports adaptability across diverse biomedical scenarios. Future improvements in accuracy may be achieved through fine-tuning techniques and ongoing advancements in LLM technologies.

## Introduction

Large-scale clinical databases are essential for healthcare operations, research, and evidence-based decision-making^1,2^. Examples of these databases include COSMIC^3^, with a pool of data from over 29,000 scientific publications; the National Cancer Database^4^, with over 34 million records of patients in the United States; or TDBRAIN^5^, a data collection from more than 1,200 psychiatric patients. The exponential growth and format heterogeneity of clinical data demand innovative approaches to efficiently manage high-volume datasets.

The Ontario Hereditary Cancer Research Network (OHCRN)^6^ is an example of such a clinical database. OHCRN is a province-wide program for individuals with hereditary cancer syndromes in Ontario. Hereditary cancer accounts for approximately 10% of all cancer cases^7^ and presents a unique opportunity for data sharing and research to improve early detection, biological understanding, and treatment. By centralizing clinical and germline genomic data and making them accessible to researchers, OHCRN aims to facilitate cancer research and connect participants with researchers and clinical trials. A significant challenge lies in the manual curation of germline genetic testing reports, which are provided as text documents in a variety of formats conforming to individual laboratory practices. This manual extraction of the relevant data points and their transformation into structured data is time-consuming, expensive and prone to human errors (e.g., typographical errors). As a representative example of multiple large databases used in biomedical research and clinical operations, OHCRN would benefit from a computational tool for extracting data from unstructured sources, which could also be applied to similar initiatives.

Traditional automated extraction tools have design and setup limitations^8^. For example, clinical data extraction for the MSK-CHORD^9^ project employed multiple Natural Language Processing transformer models that required individual training on different curated data sets, plus additional rule-based models to process more structured data. Recently, Large Language Models (LLMs) have made great strides in improving both the power and ease of use for data extraction applications. In the biomedical domain, BioBERT^10^, MT-clinical BERT^11^, and Med-PaLM^12^ demonstrate capabilities in entity extraction and normalization. Modern foundation models, such as GPT^13^, LLAMA^14^ and Claude^15^ offer unstructured data extraction via in-context learning. For instance, GL-NER^16^ employs a few-shot approach and proposes a Universal Prompt template to improve the model’s Named Entity Recognition (NER) tasks. By contrast, GPT-NER^17^ re-frames the NER task from sequence labelling to text generation, making it easier for LLMs to recognize entities while still following the general paradigm of in-context learning.

Furthermore, genetic testing reports present unique challenges for data extraction such as intricate tables, genetic variants nomenclature and highly specialized terminology, and the presence of established or novel variants. Format also varies across laboratories, requiring deep contextual understanding to correctly interpret findings.

To address these limitations, we propose LEI (LLM-based Extraction of Information), an integrated framework that leverages computer vision and LLMs to extract healthcare data from non-standardized, unstructured text sources, and provide machine-readable output suitable for database curation. The primary purpose of LEI is to streamline the data extraction and curation process of molecular lab reports, empowering data curators to process reports at a much accelerated pace and comparable accuracy.

## Methods

As part of the OHCRN initiative, germline genetic testing results for at-risk individuals undergoing testing in one of nine provincially funded laboratories will be collected from 20 genetics clinics across Ontario. Data will be centralized within a central repository (Supplementary Figure 1). The lab reports must be processed manually by OHCRN’s data curators, who locate and extract the information to be entered into the OHCRN data repository in accordance with its data model (Supplementary Figures 2A, 2B).

### The LEI data extraction framework

LEI consists of two modules: a computer vision module for converting physical documents into digital text, and a data extraction module, which employs multiple strategies, such as regular expression searches and LLM-based entity recognition to extract genetic testing data. LEI’s output is represented as JSON-formatted key-value pairs (Figure 1). To address the unique characteristics of germline genetic test reports, LLM prompts were designed with the support of an expert curator and LEI employs multiple standard nomenclatures for the definition of regular expressions and the trie structure.

**Figure 1.**
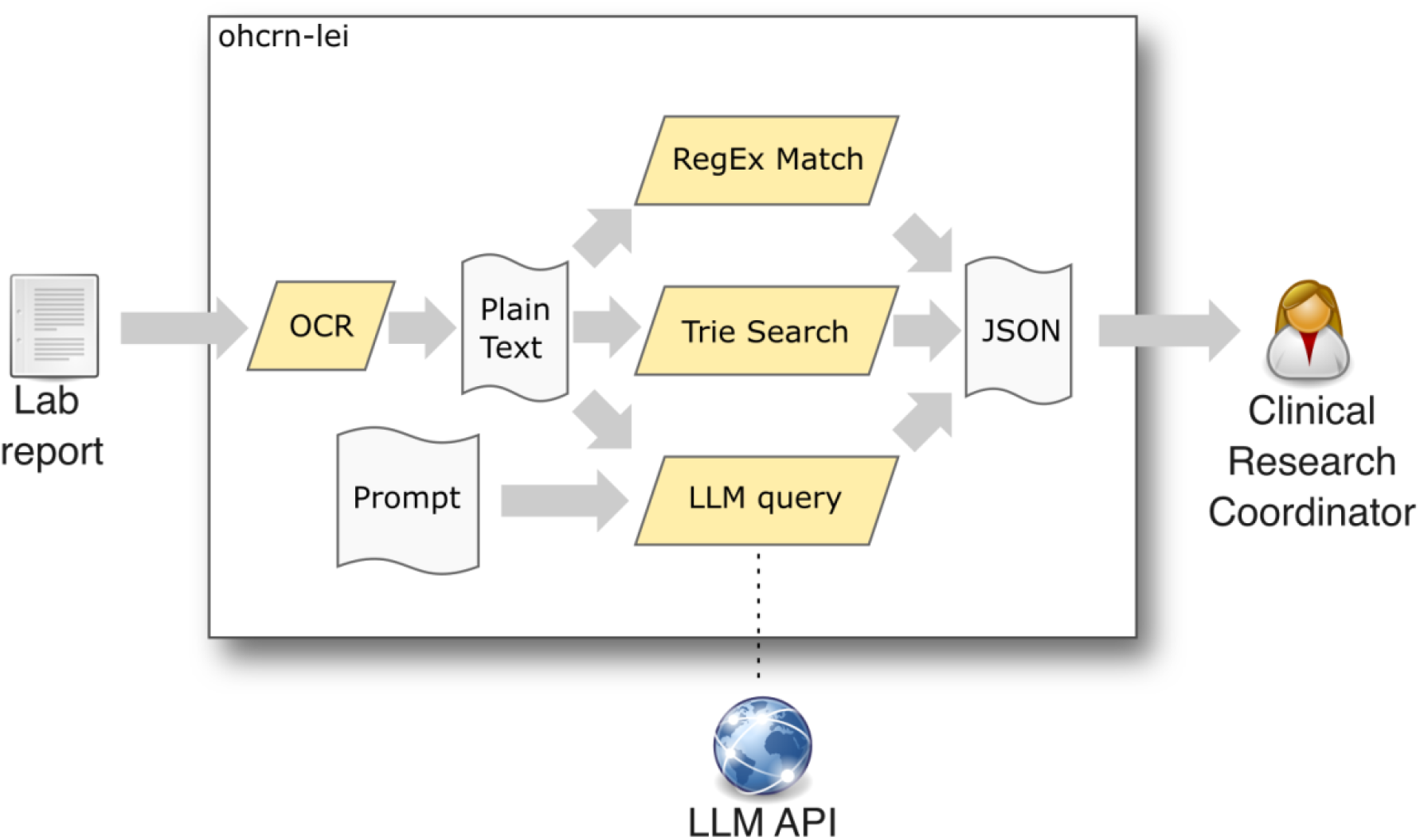
Overview of LEI. Lab reports are digitized using the OCR tool, and the text is then passed to the LLM, RegEx and Trie Search. The LLM also takes as input the prompts defining the entities to extract and a system message that guides the model’

### The computer vision module

The computer vision module accepts scanned or photographed documents in PDF or JPG format and converts them to digital text. When designing this module, multiple optical character recognition (OCR) libraries were considered, namely Tesseract^18^, EasyOCR^19^ and Amazon Textract^20^. A comparative analysis by Vedhaviyassh and colleagues found that EasyOCR outperforms Tesseract OCR accuracy by 5% due to its deep learning capabilities for object recognition^21^. Textract was discarded for not being open-sourced and the EasyOCR library was chosen instead. The LEI computer vision module was implemented in Python.

### The data extraction module

The data extraction module takes the digital text output from the computer vision model and extracts relevant biomedical data, such as clinical report types, dates, and descriptors of identified genomic variants. Extraction was performed using a combination of pattern matching with regular expressions, Trie searches, and zero-shot LLM prompting.

Pre-defined regular expressions for a number of biological entity identifiers were obtained from the following sources: patterns for variant identifiers in HGVS format^22^ from the hgvs-regexp repository^23^ for the DNA level and mavehgvs^24^ for the protein level nomenclature. Accession patterns for variant accessions and cross references from OMIM^25^, ClinVar^26^, dbSNP^27^, COSMIC^3^, ClinGen^28^, and UniProt^29^ were obtained from their respective authoritative sources. Finally, chromosome identifier patterns were created manually.

To recognize official gene symbols and aliases as defined by the HUGO human gene nomenclature committee (HGNC)^30^, a Trie Search algorithm^31^ was implemented. The HGNC gene definitions file was downloaded and all symbols and aliases were compiled into a prefix tree structure. This structure was then used as a finite state machine to step through text documents, identifying all matching substrings.

For the LLM component, OpenAI GPT-4o^32^ was used, as it has previously been demonstrated to be highly accurate and flexible, enhancing its ability to comprehend and generate cross-modal content^33^. Access to GPT-4o was handled via the Chat Completions API^34^, which provides two different communication roles: a system message role to set the expected behavior from the model and a user message role to provide requests to which the assistant will respond^35^. The latter was used to input text from the molecular reports, from where the data was to be extracted.

To ensure consistency in the LLM’s response when faced with the same input text, a softmax temperature parameter of zero was used. This value favors deterministic and highly probable outputs^36^.

A prompt-engineering paradigm described by Zamfirescu-Pereira et al.^37^ was used, which describes three relevant strategies:

1. desired interactions: examples for fields that had specific formats or needed additional context to be found by the LLM, such as identifiers;
2. code-like or template-based prompts: prompts using Markdown syntax and
3. repeated command prompts: prompts in which repeated commands were inserted for the cases when the resulting data from the extraction pipeline was missing information or when the LLM ignored commands initially present at just one point in the input prompt.

A Python application for LEI was provided to run the regular expression matching, connect to the OpenAI API, process the prompt and input text, retrieve the result and merge the final JSON output. Prompts for common extraction tasks were packaged with the application, while new custom prompts were provided as plain text files.

### Evaluation method

Nine PDF test files containing scanned de-identified molecular lab reports were used, each from a different clinical laboratory in Ontario, covering a variety of gene panels. The reports varied in length from 2 to 8 pages. Each had a different structure, depending on the issuing institution. To differentiate between the performance of the OCR component and the data extraction component, manual plain-text conversions of the documents that could be used to bypass the OCR were also created. To create a gold standard extraction benchmark, manual extractions were performed multiple times on all nine documents by two expert curators and the results reconciled.

Focus was placed on the evaluation of three different extraction tasks: “Report”, “Molecular Test”, and “Variant”, each of which extracted a different set of entities (Supplementary Table 1). A Python script was used to normalize and compare the JSON output against the gold standard and calculate the number of true positive, false positive and false negative entries.

The expected time savings afforded by LEI was quantified by measuring the time needed to perform the Report, Molecular Test and Variant extraction tasks across the nine molecular reports by LEI, as well as by a human curator performing the task manually.

## Results

LEI was capable of performing the extraction ~2 times faster than the manual process. The mean extraction time by LEI, including the time needed to review results, was 7.59 minutes, while the mean manual extraction time by the human curator was 14.88 minutes (Figure 2A).

**Figure 2.**
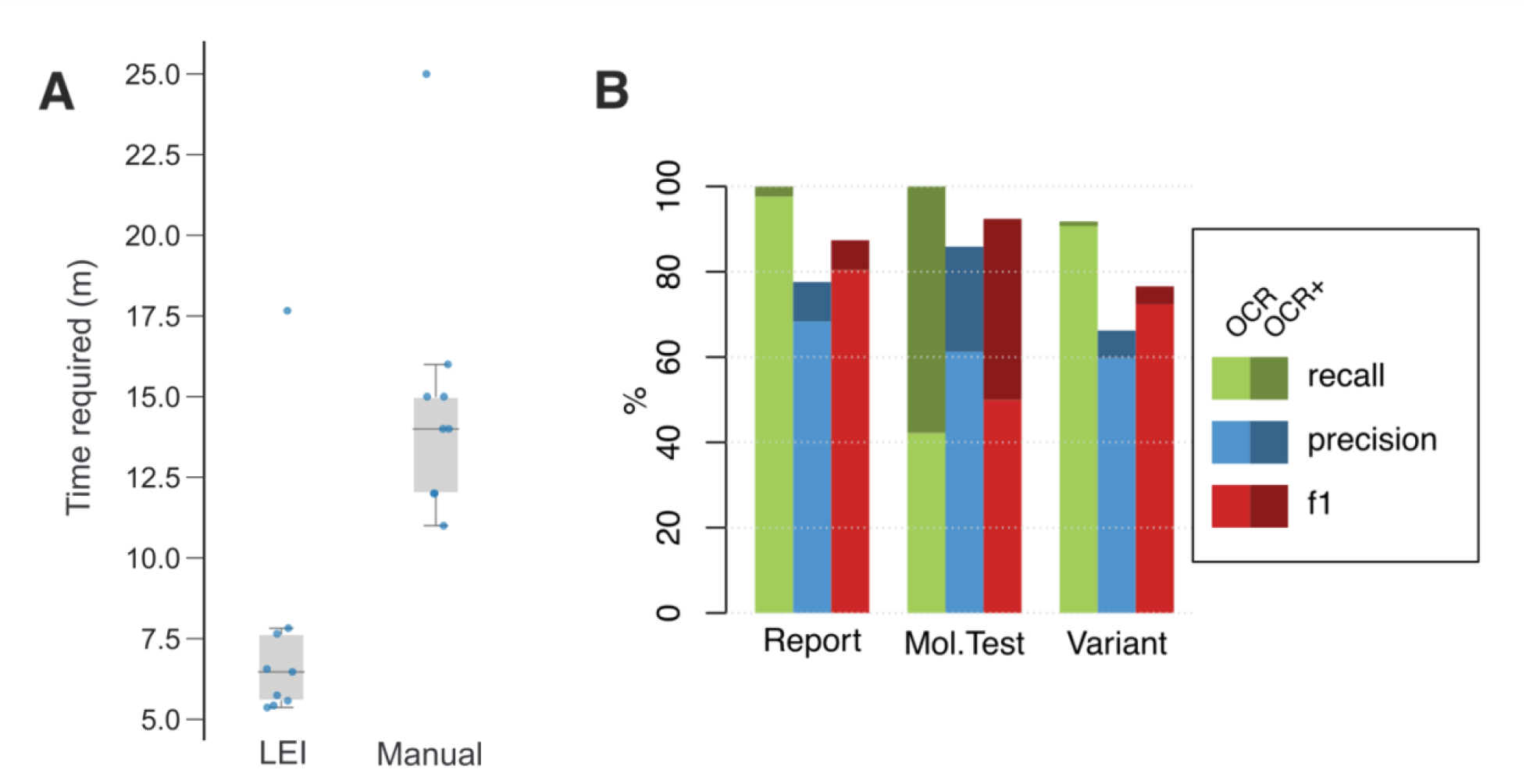
Extraction results. **(A)** Time taken to extract the information from all reports, using LEI vs manual extraction. LEI times include the time required to review results by expert curator **(B)** Precision, recall and F1-score for each extraction task with and without OCR errors. Light colors show the first scenario, which included OCR errors produced by the quality of scanned documents. Dark colors show the second scenario, in which the OCR errors were manually corrected to ensure input text quality

### Extraction accuracy

To evaluate the reliability of the results, the automatic extractions were compared against the manually curated gold standard, determining true positives, false positives, and false negatives for each entity type. True positives represent data that is correctly extracted and accurately mapped to the appropriate OHCRN data model field. Conversely, false positives are extracted values that are incorrectly mapped or are not present in the original text, akin to “hallucinations.” False negatives are missed entities that should have been extracted, effectively representing “missing values.” We then calculated precision, recall, and F1-scores. Precision is the ratio of true positives to all extracted positives. Recall measures the ratio of true positives to all actual entities meant for extraction. Finally, the F1-score provides a harmonic mean of precision and recall.

Errors can originate from the OCR process or the information extraction process; consequently performance metrics were measured in two different scenarios. The first scenario considers both the OCR and the extraction processes together. In the second scenario, OCR issues were manually corrected to ensure accurate text input for extraction. In this scenario, the Report extraction task successfully achieved the desired score range, with an F1 of 87.4%. The Molecular Test task achieved an F1-score of 92.5, with 100% recall. Notably, the use of OCR had the biggest impact here. Lastly, the Variant task showed an F1-score of 76.6%, explained by a high recall of 90.7% and an acceptable precision of 66.2% (Figure 2B).

Diving deeper into the performance metrics, the “Ordering Clinic” for the “Report” task presented the largest hurdle. The LLM used the testing lab name instead since this value was not included in most reports, generating false positives. For “Molecular Test”, false positives stemmed from the Trie search identifying words identical to real gene names. Besides, OCR issues caused many gene symbols to be missed, particularly in a single report with a continuous list of over 800 gene symbols. The “Sample type” recall suffered as most values are implicit in the reports and require further genetics expertise to deduce them. Finally, the “Variant” task precision was affected by false positives in the “variation code” and “coding HGVS” entity types. The former was caused by persistent hallucinations of the LLM, while the latter was caused by regular expressions incorrectly extracting variants mentioned in the report that are not the targeted data, i.e., the variant identified in the patient (Supplementary Figure 3).

## Discussion

LEI is an adaptable framework that does not require set-up, pre-training or curated data, which can also be extended to new tasks described in natural language. These features render LEI accessible to clinical researchers with minimal computational experience. With an overall precision exceeding 80% and over 2-fold time savings, LEI enables genetic testing data extraction from unstructured texts in a standardized, accurate and fast manner. LEI replaces the manual extraction subprocess (Supplementary Figure 2C) and assists curators in the validation step. This enables curators to review and update if needed to ensure the quality and correctness of the data.

We demonstrate how LEI improves the efficiency of the data extraction process for lab reports that are part of the OHCRN database. LEI accurately extracts genetic testing report data by combining the LLM capabilities to process unstructured texts with the ability of Trie search and regular expressions to identify data unique to genetic testing. Thanks to the remote API calls, LEI’s runtime is generally independent of local hardware and can thus be run on a low-end laptop.

Recent similar approaches on biomedical information extraction show precision, recall, and F1-scores between 70% and 90%^11,16,38–40^ (Supplementary Table 2). Even if an automated extraction system does not reach perfect accuracy, having a recall of e.g., 70% means the curator will have to manually process the remaining 30% of the total data, which is easier than processing all the information^41^. With LEI, the burden of manual extraction was reduced for the OHCRN clinical coordinator, reducing the time needed for this recurrent process and eliminating the chance of human data input errors when sending information to OHCRN’s repository.

The precision-recall analysis of LEI’s output showed its quality to be on par with, and even surpass, other existing LLM-based extraction methods. In all tasks, LEI exceeds the established F1-score threshold of 70%. LEI’s extraction accuracy also surpasses manual extraction by humans. Mathes et al.^42^ assessed the frequency of data extraction errors in manual systematic reviews and found error rates could reach up to 50%. This fact underscores the importance of building a gold standard benchmark through consensus between multiple manual extraction passes performed by multiple curators that could be used to validate LEI’s results.

While most errors were caused by incorrect extractions by the LLM, a considerable number stemmed from OCR problems, where the quality of scans hindered character recognition. OCR accuracy varied significantly across documents, influenced by scan quality, typefaces, and formatting. The variability caused by these errors propagated through our pipeline, especially when leveraging LLMs to extract relevant information. OCR also commonly struggled with recognizing certain variant information, such as protein and coding changes, resulting in incomplete data extraction.

LEI provides a simple interface with pre-configured extraction tasks as well as support for defining new tasks using natural language with a few formatting rules. This enables researchers or clinicians with few or no programming experience to easily direct LEI’s results, and opens up the possibility of using the solution in multiple contexts with minimal effort. The sole use of LLMs for data extraction would be ideal for simplifying the pipeline even further, as developing regular expressions can be intimidating or unapproachable without specific expertise^43^, which can be the case for the average researcher or clinician. Our pipeline could then be improved by trading the regexes for LLM-prompted extraction.

While our current data extraction pipeline demonstrates significant advantages in time efficiency and accuracy for extracting relevant information from unstructured healthcare documents, certain limitations need to be acknowledged. These limitations are areas for possible future improvements.

LEI was developed using a limited set of reports from nine molecular testing laboratories across Ontario. The manual extraction performed by the expert curators ensures an effective gold standard benchmark for validation. However, the limited number of sample cases available for testing means that our evaluation may not account for the full variability of report formats and structures across institutions. As the OHCRN initiative develops, more reports will become available for a larger test dataset, thus enabling more extensive evaluation. In combination with feedback from clinical data curators, these new reports can be used to continuously improve LEI.

A major concern regarding all current LLMs is hallucinations, i.e., the presence of false assertions in the generated responses of the model^44^. In earlier iterations of LEI, hallucinations were observed, especially for extracted variant descriptors, where the LLM produced results that complied with the descriptor format, but with values that were absent in the reports. To avoid hallucinations, regular expressions were used for fields with standardized formats. Additionally, LEI’s implementation of the prompting strategies mentioned by Zamfirescu-Pereira et al.^37^ proved to be effective for the data extraction task.

LEI relies on GPT-4o as the primary LLM. However, open-source and domain-specific models offer promising alternatives. These models provide flexibility to customize and adapt LEI for specific healthcare contexts. A significant advantage of open-source models is their ability to be deployed locally, eliminating the need to transmit sensitive patient data to third-party servers and thereby reducing privacy concerns. Local deployment can also increase LEI’s adoption by other institutions with similar interests to OHCRN, allowing them to confidently implement our pipeline whilst retaining full control over the data and maintaining patient anonymity.

Open-source fine-tuning tools such as Unsloth^45^ can be integrated into LEI, offering a customizable and efficient framework for fine-tuning LLMs to better suit data extraction tasks. However, a recent study by Dorfner et al.^46^ found that biomedical-specific LLMs often underperform compared to generalist models on tasks involving unseen medical data. Yet, the study highlights the use of retrieval-augmented generation (RAG) to combine the strengths of general purpose models with external domain-specific knowledge bases. Incorporating RAG can enable an LLM to extract relevant information more accurately without sacrificing general performance capabilities. Combining fine-tuning and RAG techniques could enhance LEI’s performance, particularly for tasks requiring specialized knowledge, such as the Variant task.

## Conclusions

In summary, we have shown that LEI provides useful assistance to clinical database human curators in terms of time savings and extraction accuracy. While it will not replace the curators, it will assist them by automatically extracting data for their review, reducing the time required for the process and reducing possible human errors. Its user-friendly, yet flexible, implementation allows for simple expansion into additional data extraction tasks as needed. While a number of limitations still exist, future iterations may soon be able to address them thanks to the rapid pace of progress in the machine learning field.

## Data Availability

The LEI application, as well as the code used for evaluation, is available at https://github.com/courtotlab/ohcrn_lei. It can be used and shared freely under the terms of the GNU Public License (GPL) 3.0.
The sample molecular lab reports used in this study were obtained from third-party laboratories with the support of the Ontario Hereditary Cancer Research Network (OHCRN). The reports were de-identified or modified using simulated but structurally representative data prior to sharing with this study. The authors do not have permission from these laboratories to share the original reports publicly. As such, the molecular lab reports cannot be made available for public access. Researchers interested in accessing similar data are encouraged to contact the respective laboratories directly to inquire about data access permissions.

## Data availability

The LEI application, as well as the code used for evaluation, is available at https://github.com/courtotlab/ohcrn_lei. It can be used and shared freely under the terms of the GNU Public License (GPL) 3.0.

## Acknowledgements

We gratefully acknowledge the contributions of the following laboratories for providing sample laboratory reports that were essential to this study: Children’s Hospital of Eastern Ontario, Hamilton Health Sciences, Kingston Health Sciences Centre, London Health Sciences, Sinai Health System, North York General Hospital, Hospital for Sick Children, Trillium Health Partners, University Health Network. Their efforts and collaboration were critical to the success of this work.

This study is conducted with the support of the Ontario Institute for Cancer Research through funding provided by the Government of Ontario.

