## Supplemental tables and figures for "LLM-based data extraction for a large cancer registry, the Ontario Hereditary Cancer Research Network"

#### – Supplementary Materials –

#### Supplementary Tables

| Report | Molecular test | Variant |
|---|---|---|
| Report date | Sequencing scope | Gene symbol |
| Report type | Tested genes | Transcript ID |
| Testing context | Sample type | Exon |
| Ordering clinic | Analysis type | Reference genome |
| Testing lab | | Variant ID |
| | | Chromosome |
| | | Coding HGVS |
| | | Genomic HGVS |
| | | Protein HGVS |

**Supplementary Table 1. Entities to extract from reports by task.** Each task corresponds to one table in the OHCRN data model. Each one of the entities is defined in the prompts given as input to the LLM.

| Author | Model | Precision (%) | Recall (%) | F1-score (%) |
|---|---|---|---|---|
| Xu et al. | TwoStepChat - zero shot | 82.33 | 88.50 | 85.31 |
| | TwoStepChat - 6 few shot | 85.94 | 89.10 | 87.49 |
| | TwoStepChat - 10 few shot | 87.35 | 91.35 | 89.31 |

| | | | | |
|---|---|---|---|---|
| | TwoStepChat - 20 few shot | 88.59 | 90.20 | 89.39 |
| Sung et al. | BERN2 on Gene/protein data | N/A | N/A | 83.7 |
| | BERN2 on Mutation data | N/A | N/A | 93.7 |
| Mulyar et al. | MT-Clinical BERT on i2b2-2010 data | N/A | N/A | 89.5 |
| | MT-Clinical BERT on i2b2-2012 data | N/A | N/A | 84.1 |
| | MT-Clinical BERT on i2b2-2014 data | N/A | N/A | 91.9 |
| | MT-Clinical BERT on quaero-2014 | N/A | N/A | 49.1 |
| Jantscher et al. | Fine-tuned BERT - high score with random sampling | N/A | N/A | ~60 |
| | Fine-tuned BERT - high score with strategic sampling | N/A | N/A | ~78 |
| Zhu et al. | GL-NER on GENIA, 10 few-shot | N/A | N/A | 33.16 |
| | GL-NER on GENIA, 50 few-shot | N/A | N/A | 43.92 |
| | GL-NER on GENIA, 100 few-shot | N/A | N/A | 44.33 |
| **LEI** | **Report task** | **77.7** | **100** | **87.4** |

| | | | | |
|---|---|---|---|---|
| | **Molecular test task** | **85.9** | **100** | **92.4** |
| | **Variant task** | **66.2** | **90.7** | **76.6** |

**Supplementary Table 2. Precision, Recall and F1-score of similar biomedical information extraction tools and LEI**

### Supplementary Figures

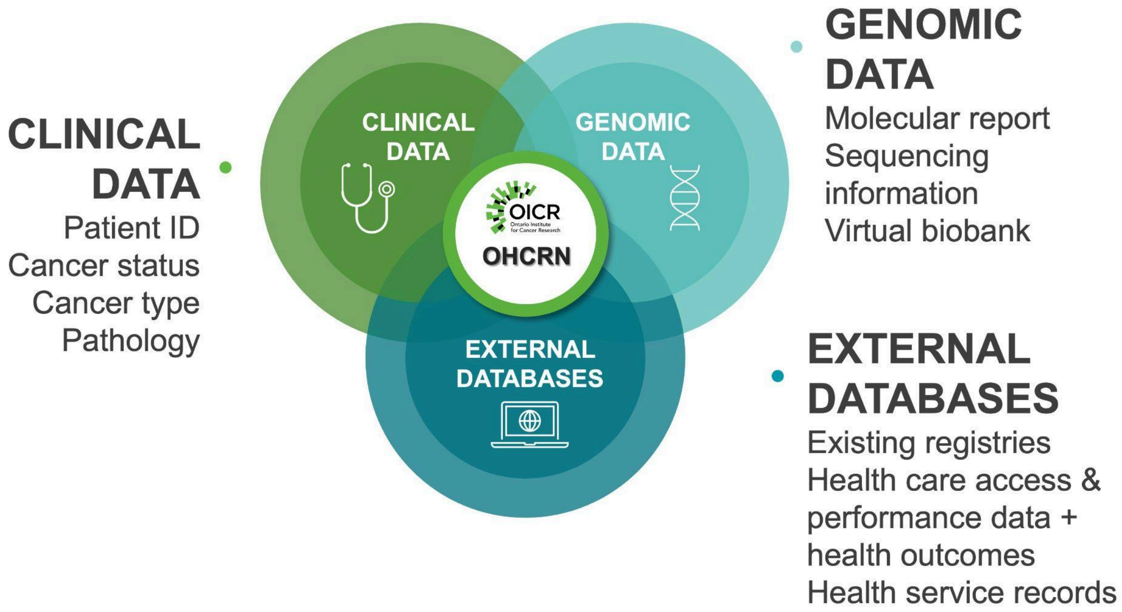


**Supplementary Figure 1. OHCRN's data sources.** Clinical information from the participant and genomic data from clinics and laboratories will be collected. Linkage with external databases will provide additional data points.

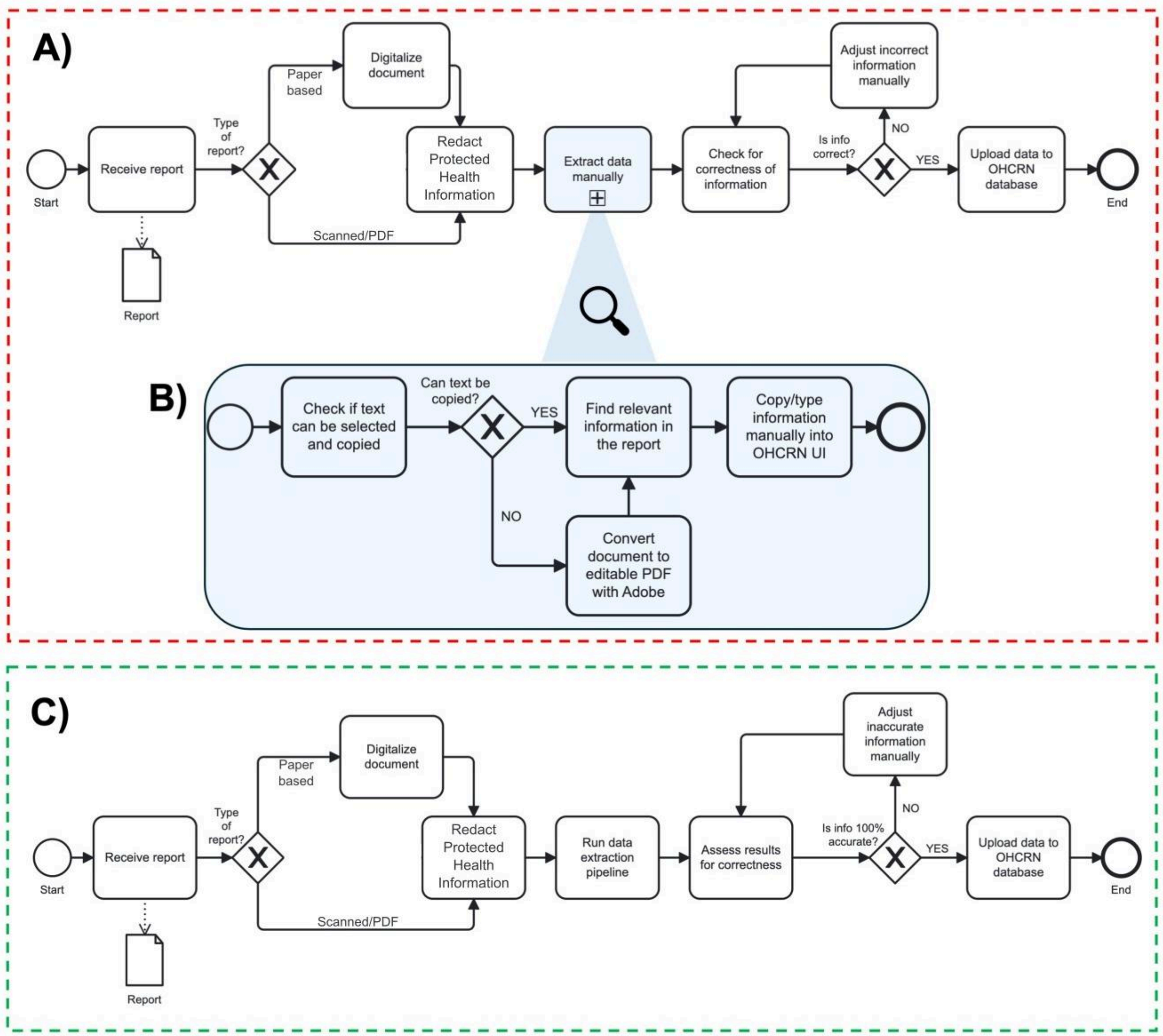


**Supplementary Figure 2. Current and proposed data extraction processes for OHCRN.** A) Current process done by OHCRN's data curator from reception of reports to upload of extracted data into OHCRN database. B) Subprocess showing the steps to manually extract information from a lab report. C) Proposed data extraction process using LEI. The manual extraction subprocess is replaced by running the pipeline.

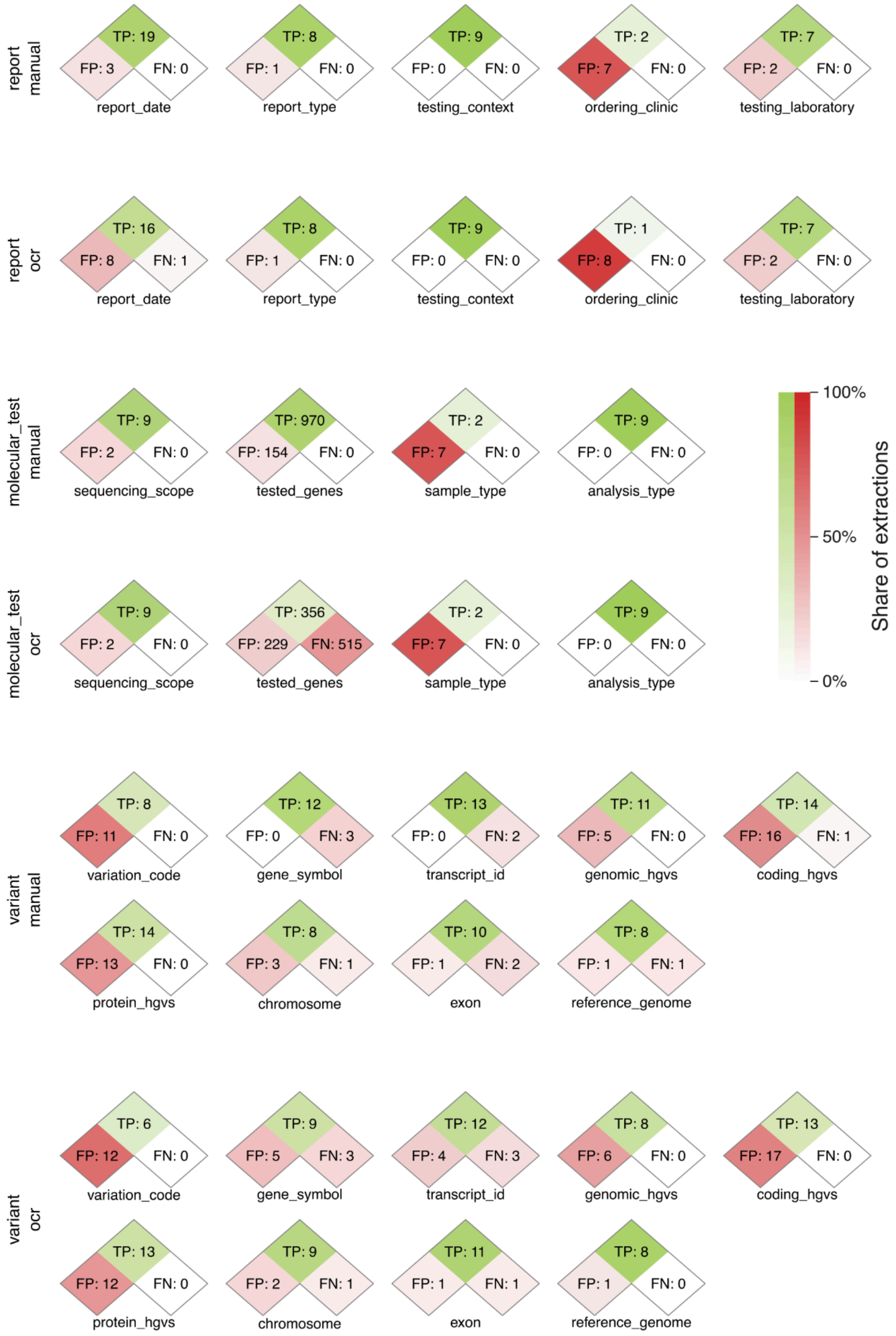


**Supplementary Figure 3. Confusion matrices for entity extractions in the Report, Variant and Molecular Test tasks, with and without OCR errors.**
